# Adult renal tubular organoids can be produced from different human individuals in a completely same protocol

**DOI:** 10.1101/2024.08.11.24311846

**Authors:** Makiko Mori, Yutaro Mori, Nakao Yuki, Shintaro Mandai, Tamami Fujiki, Hiroaki Kikuchi, Fumiaki Ando, Koichiro Susa, Takayasu Mori, Yuma Waseda, Soichiro Yoshida, Yasuhisa Fujii, Eisei Sohara, Shinichi Uchida

**Affiliations:** Department of Nephrology, Graduate School of Medical and Dental Sciences, Tokyo Medical and Dental University, Tokyo 113-8510, Japan; Department of Urology, Graduate School of Medical and Dental Sciences, Tokyo Medical and Dental University, Tokyo 113-8510, Japan

**Author notes:** **Lead contact and Corresponding author:** Yutaro Mori, MD, PhD, Department of Nephrology, Graduate School of Medical and Dental Sciences, Tokyo Medical and Dental University, 1-5-45, Yushima, Bunkyo-ku, Tokyo 113-8510, Japan. These authors contributed equally to this work (Makiko Mori, Yutaro Mori).

**Keywords:** organoids, tubuloids, pathological model, chronic kidney disease

## Abstract

**Introduction:** Organoids are miniature organs produced by newly emerging technologies. Kidney organoids originated from human inducible pluripotent stem cells (iPSCs) were developed to recapitulate renal diseases. However, producing iPSC kidney organoids from multiple individuals at the same time and in a uniform condition is still impossible. Here, we report adult renal tubular organoids, “tubuloids”, established from primary renal epithelial cells from multiple human individuals in a uniform manner.

**Methods:** Kidneys obtained from patients due to the surgery for malignancy were minced into small pieces, and primary renal epithelial tubule cells are cultured. 4 patients had normal kidney function and 4 had mild chronic kidney disease (CKD). Growth factors were added to the primary cultured cells at the same time and Matrigel was added to these 8 lines.

**Results:** Primary cultured renal epithelial cells from normal kidneys showed a large number of fine, swollen epithelial appearance. On the other hand, primary cultured kidney epithelial cells from mild CKD kidneys were smaller and slightly elongated than those of normal kidneys. The growth speed was faster in normal kidney cells than in mild CKD cells. At the beginning of the three-dimensionalization (day 0), normal renal tubuloids grew faster than mild CKD tubuloids. The difference in size between normal tubuloids and mild CKD ones became less noticeable on day 5. Both types of tubuloids reached almost same size on day 10. All 8 strains are of different human origin, and uniform tubuloids could be produced at the same time and in a uniform protocol.

**Conclusion:** In terms of pathological models, the differences between mouse models and humans cannot be ignored, and there is a great need for a more human-like model of human pathology from both medical and research perspectives. Our renal tubular organoids can be produced in a uniform manner at the same time. It is expected to be used as a new type of convenient human pathological model.

## Introduction

Biomedical engineering has made remarkable progress in recent years, and multiple diseases of various organs have been elucidated by using organoids which are miniature organ-like structures produced by newly emerging technologies and serve as an alternative and more human-like pathological model [1, 2]. Chronic kidney disease (CKD) is an irreversible, progressive disease characterized by glomerular sclerosis, tubular atrophy and collapse, and fibrosis and infiltration of inflammatory cells in the stromal cells [3]. However, the pathogenesis of CKD is not yet fully understood. No curative therapy has been developed as well. There is a great need to understand CKD from both research and medical perspectives.

One of the reasons for the delay in CKD research may be the fact that kidney disease is largely influenced by aging [4]. Rats and mice are mainly used for recapitulating kidney diseases. However, they cannot accurately reflect the aging of human organs that are over 80 years old. Kidney organoids established from human inducible pluripotent stem cells (iPSCs) have emerged as a new pathological model to replace rodents [5, 6]. At the present, however, iPSC kidney organoids have not been succeeded to reproducing kidney condition later than the embryonic stage [7]. It is difficult to reflect the aging and senescence process. Another problem with iPSC kidney organoids is that they require extremely skilled technique to fabricate them.

When focusing on pathophysiology of CKD, the most serious problem of iPSC kidney organoids is that they cannot be created from different individuals in a completely uniform manner. To make iPSC kidney organoids, we need to start to establish nephron progenitor cells (NPCs) from iPSCs [5, 6]. Culture condition especially for inducing differentiation into NPCs varies greatly depending on the origin of the iPSCs. Furthermore, NPCs do not necessarily become kidney organoids under the uniform differentiation induction conditions on different strains. This matter that iPSCs from different individuals cannot form kidney organoids with a uniform condition is a critical and fatal weakness for studying CKD having a very large diversity of causes, age, race, sex, and speed of progression. This problem makes accurate analyses of CKD and drug testing for uniform conditions almost impossible. As long as we know, iPSC kidney organoids produced from multiple human-derived iPSCs in a uniform condition has not yet been reported. In addition, even mouse models recapitulating CKD-like diseases have not succeeded in reproducing the diversity that each individual human has which was mentioned above.

Recently, CKD has been considered to be a disease headed by the renal tubules more than the glomerulus [8, 9]. We have focused on renal proximal tubules and have successfully developed tubular organoids derived from primary human renal proximal tubular epithelial cells (hRPTECs) [10–12]. In this study, we have succeeded to produce human adult renal tubular organoids, “tubuloids”, from different individuals at the same time in a uniform condition. Our strategy can provide a new insight to analyze pathophysiology of kidney diseases including CKD in a personalized or semi-personalized fashion.

## Methods

### Cell culture experiments

Human kidney samples were obtained from kidneys clinically removed from patients with renal cell carcinoma or urothelial malignancy at Tokyo Medical and Dental University Hospital. The protocol was approved by the Institutional Review Board of the Ethics Committee of Tokyo Medical and Dental University (M2022-005). Then, primary hRPTECs were cultured from non-infiltrated areas of the kidney by malignancy, removed as described above.

First, human kidney cortex was cut into small pieces and dissolved in a solution of collagenase type II (1.0 mg/mL) (Worthington Biochemical, NJ, USA). Then, Bovine Serum Albumin (Nacalai Tesque, Kyoto, Japan), Antibiotic-Antifungal (ThermoFisher Scientific, MA, USA), hydrocortisone (ThermoFisher Scientific, MA, USA), ITS liquid media supplement (Sigma-Aldrich, MO, USA), and human recombinant epidermal growth factor (EGF) (ThermoFisher Scientific, MA, USA) in hRPTEC culture medium (DMEM/F-12 (Nacalai Tesque, Kyoto, Japan). Cells were then cultured in a CO2 incubator (5% CO2) at 37°C for 5∼7 days.

### Human renal tubuloid

hRPTECs were seeded on ultra-low adhesion plates using Advanced RPMI 1640 medium with 5% fetal bovine serum (SERANA, Brandenburg, Germany). Two days later, Matrigel (Corning, NY, USA) was added, followed by the addition of Advanced RPMI with 5% FBS plus EGF, FGF (ThermoFisher Scientific, MA, USA), and HGF (ThermoFisher Scientific, MA, USA). We define these media containing growth factors above as “tubuloid media”. 1∼2 weeks of incubation in a CO2 incubator (5% CO2) at 37°C completed the tubuloids. The medium was changed about twice a week. No other work was required; basically, they should be left alone.

### Microscope observation

We observed and took pictures of all the cells and the tubuloids by using BZ-X800 (KEYENCE, Oasaka, Japan).

## Results

### We have established 8 individuals’ primary hRPTECs from resected kidneys

**Table 1** shows the detailed characteristics of 8 individuals who had nephrectomy due to mainly malignancies. 4 individuals have normal kidney function (eGFR > 60 mL/min/1.73 m^2^, named N-1 to N-4), and the other 4 have mild CKD (eGFR 45-60 mL/min/1.73 m^2^, named C-1 to C-4). We established these 8 strains of primary hRPTECs in 2-dimensional cultured condition. Interestingly, as long as we experienced, lower kidney function did not affect the efficiency of primary culture establishment.

**Table 1.**
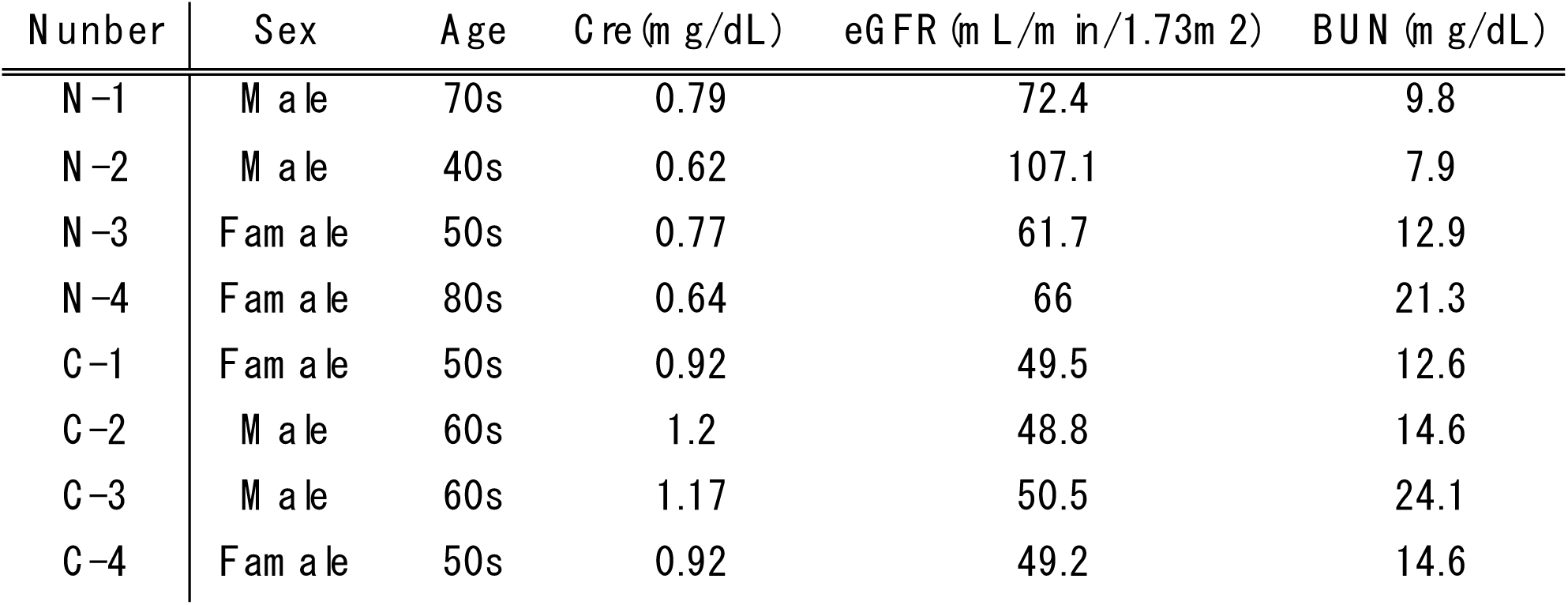
Detailed data of the study participants focusing on sex, ages, and renal functions.

### Primary hRPTECs individuals with mild CKD showed relatively elongated morphology, not typical cobblestone appearance as renal epithelia cultured cells

We observed 2-dimensional culture of all the hRPTECs in passage 3 generation. All the hRPTECs became confluent in approximately one week. Cell growth is slightly faster in normal-derived cells. The hRPTECs from individuals with normal kidney function showed cobblestone appearance **(Figure 1, the top row**). Those from individuals with mild CKD showed relatively elongated morphology (**Figure 1, the bottom row**).

**Figure 1.**
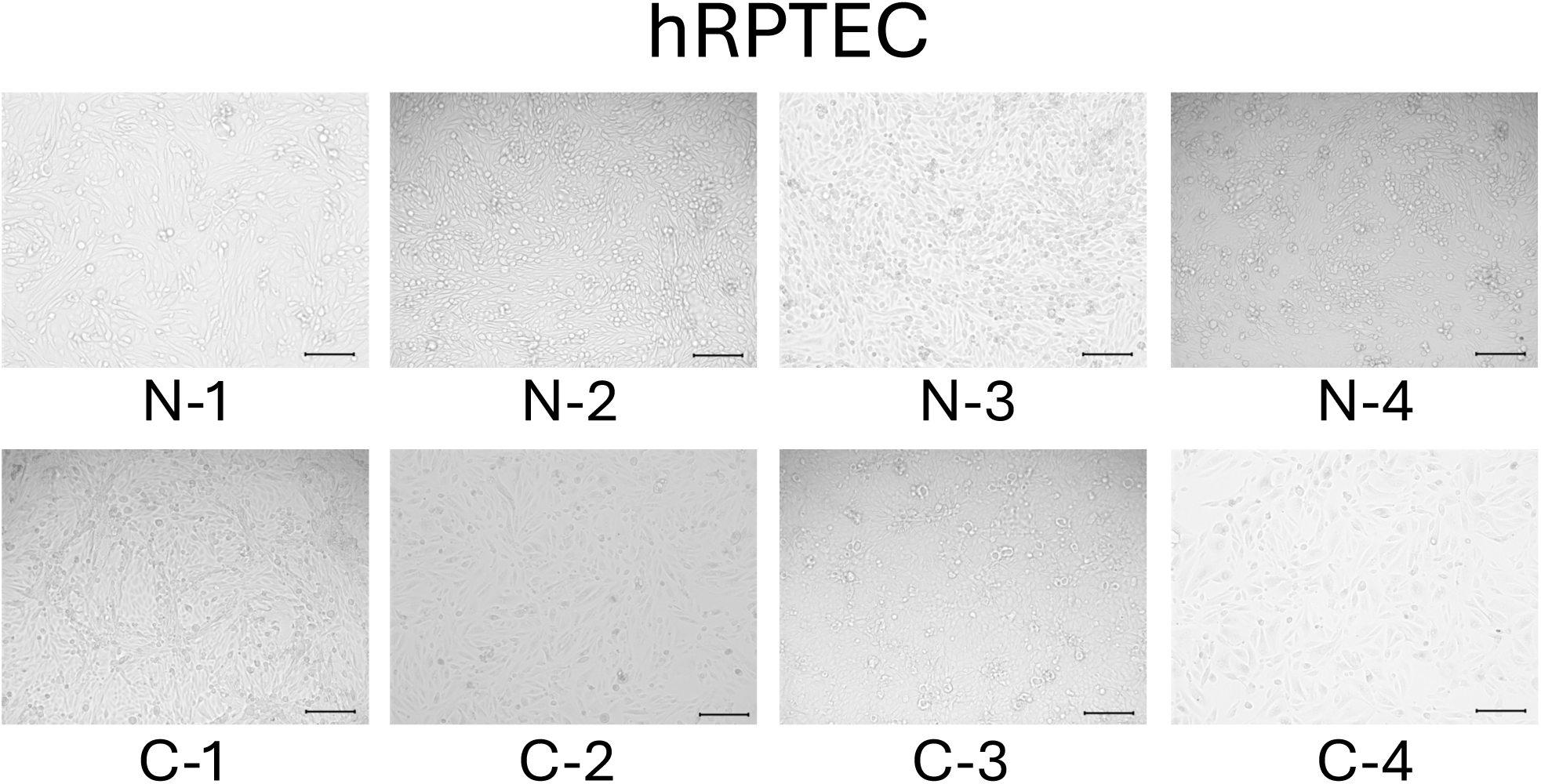
Primary hRPTECs individuals with mild CKD showed relatively elongated morphology, not typical cobblestone appearance as renal epithelia cultured cells. The top row shows cells from individuals with kidney function (N-1 to N4), and the bottom row shows those from mild CKD (C-1 to C-4). Scale bars : 200µm.

### We have succeeded to produce human adult renal tubular organoids, “tubuloids”, from different individuals at the same time in a uniform condition

Next, the hRPTECs were tridimensionalized and made into tubuloids. We seeded hRPTECs on ultra-low adherence plates with 5% FBS in advanced RPMI media. 2 days after seeding, we added Matrigel as the final concentration of it became 10%. We defined day 0 was the day on which Matrigel was added. On day 1, we added

First, we observed the tubuloids on day 4. Normal cell-derived tubuloids immediately began to form a clear, round, bubble-like structure (**Figure 2, the top row)**. In contrast, the mild CKD cell-derived tubuloids had a bumpy, small structure (**Figure 2, the bottom row**). Structures of the tubuloids on day 6 showed little change from those on day 4 (**Figure 3**).

**Figure 2.**
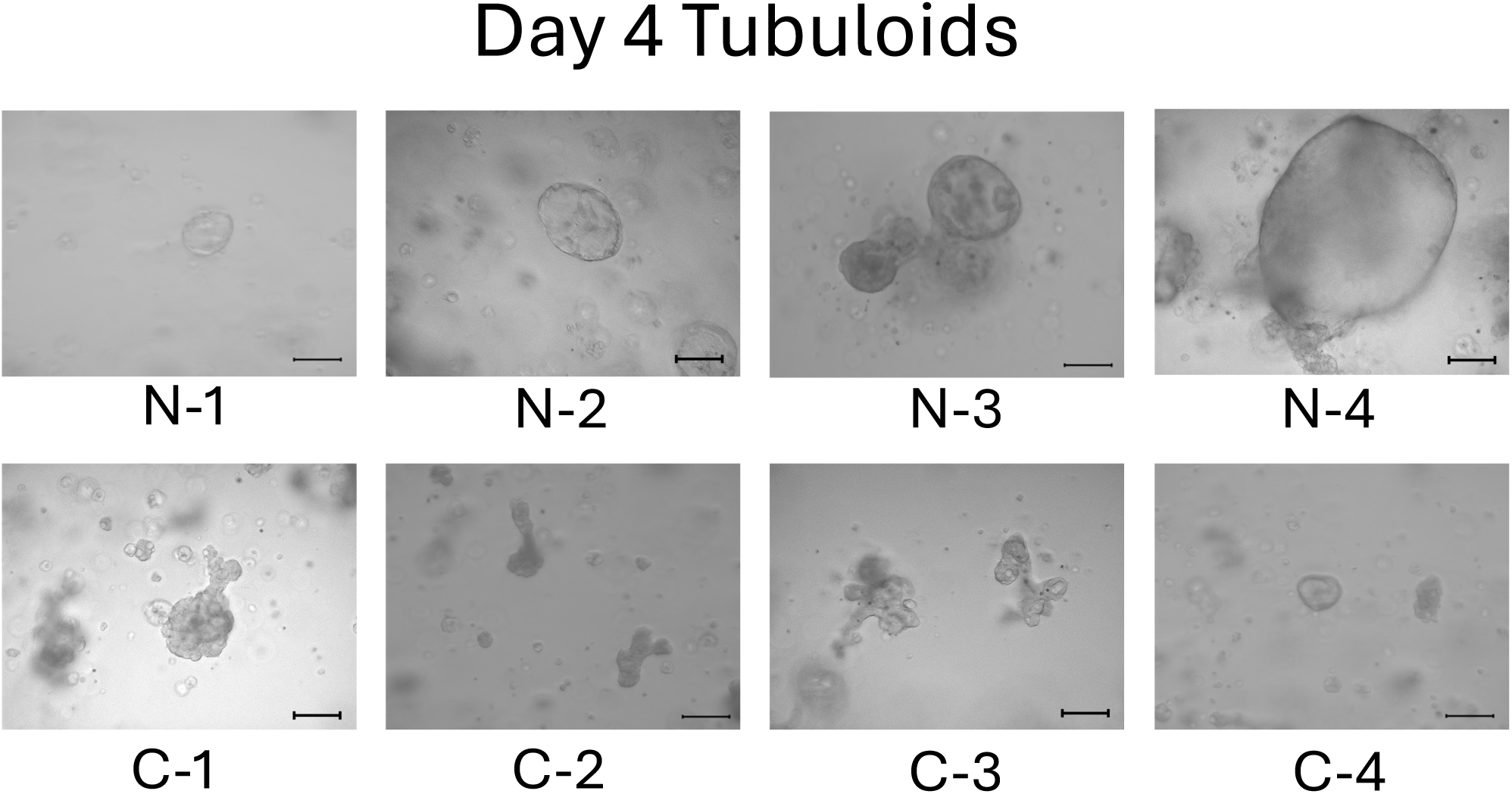
On day 4, normal cell-derived tubuloids began to form a clear round bubble-like structure, but the mild CKD cell-derived tubuloids have a structure of bumpy little things. The top row shows 3D-cultured cells from individuals with kidney function (N-1 to N4), and the bottom row shows those from mild CKD (C-1 to C-4). Scale bars : 200µm

**Figure 3.**
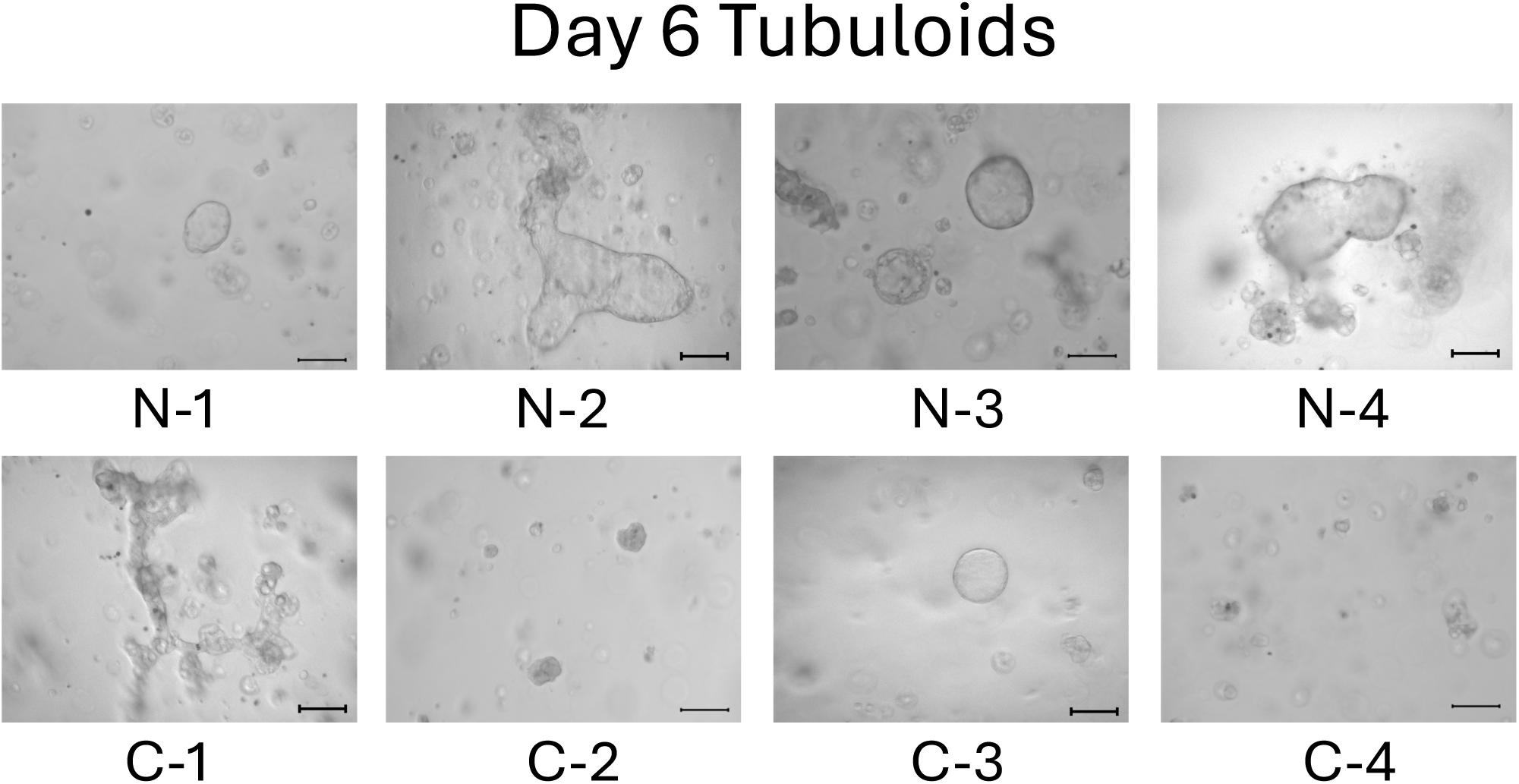
On day 6, mild CKD cell-derived tubuloids still tended not to form a bubble-like structure. The top row shows 3D-cultured cells from individuals with kidney function (N-1 to N4), and the bottom row shows those from mild CKD (C-1 to C-4). Scale bars : 200µm

On day 13, the difference between normal and mild CKD tubuloids was widened. Normal cell-derived tubuloids had beautifully large circular structures **(Figure 4, the top row)**. In contrast, the mild-CKD cell-derived tuberoids showed the small circular structures sporadically **(Figure 4, the bottom row)**. By day 18, the tubuloids with circular structures were even larger. The growth of C-3 was particularly tremendous, greatly exceeding the size of normal cell-derived tubuloids (**Figure 5**). By day 21, even the C-2 and C-4 tubuloids, which had been difficult to transform into circular tubuloids before, showed a beautiful circular structure as same as tubuloids originated from other individuals (**Figure 6**).

**Figure 4.**
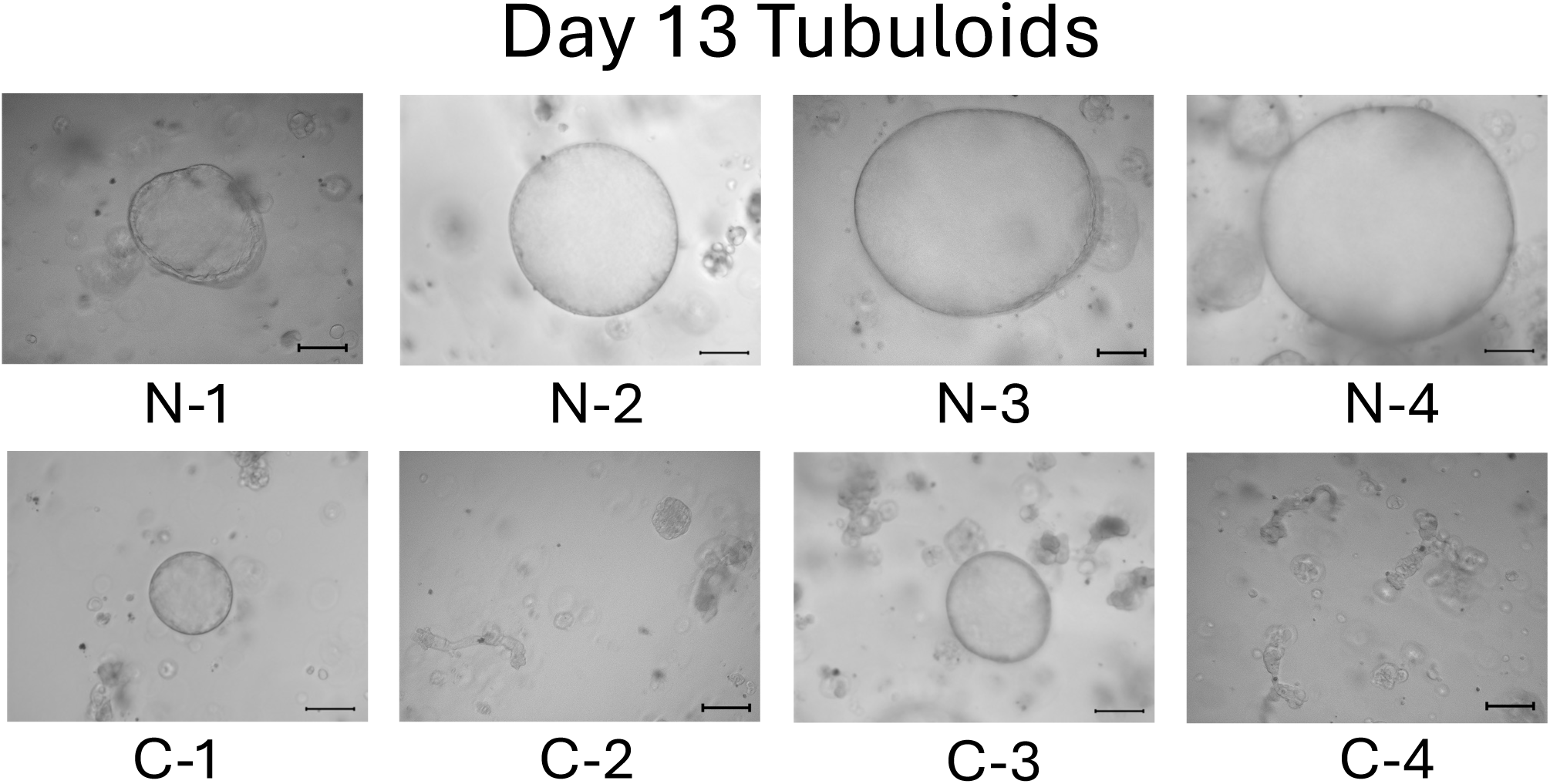
On day 13, all of normal cell-derived tubuloids formed almost completely, and some of mild CKD cell-derived tubuloids started to form a tubuloids. The top row shows 3D-cultured cells from individuals with kidney function (N-1 to N4), and the bottom row shows those from mild CKD (C-1 to C-4). Scale bars : 200µm

**Figure 5.**
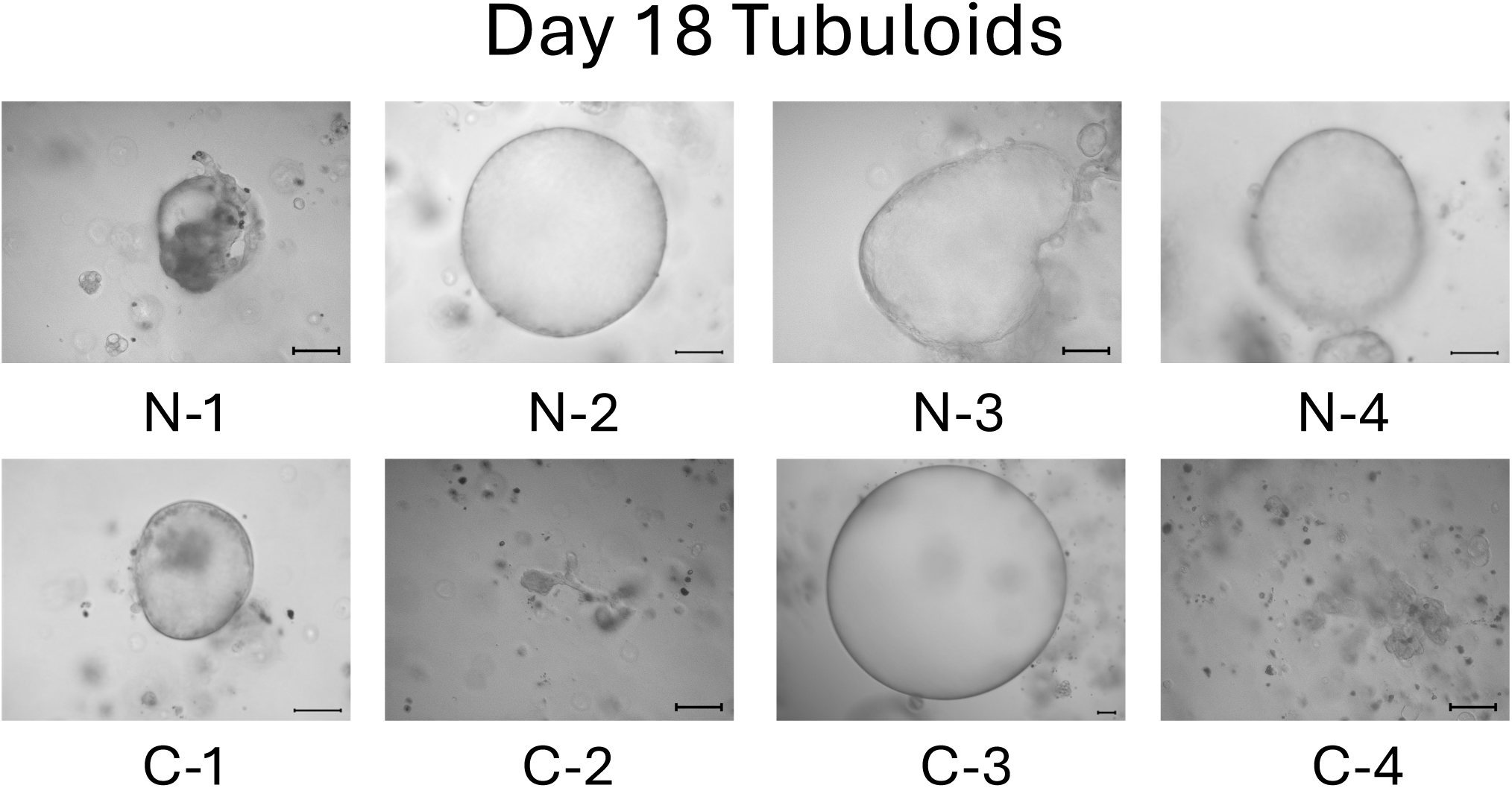
On day 18, some of mild CKD cell-derived tubuloids became larger, but others not. The top row shows 3D-cultured cells from individuals with kidney function (N-1 to N4), and the bottom row shows those from mild CKD (C-1 to C-4). Scale bars : 200µm

**Figure 6.**
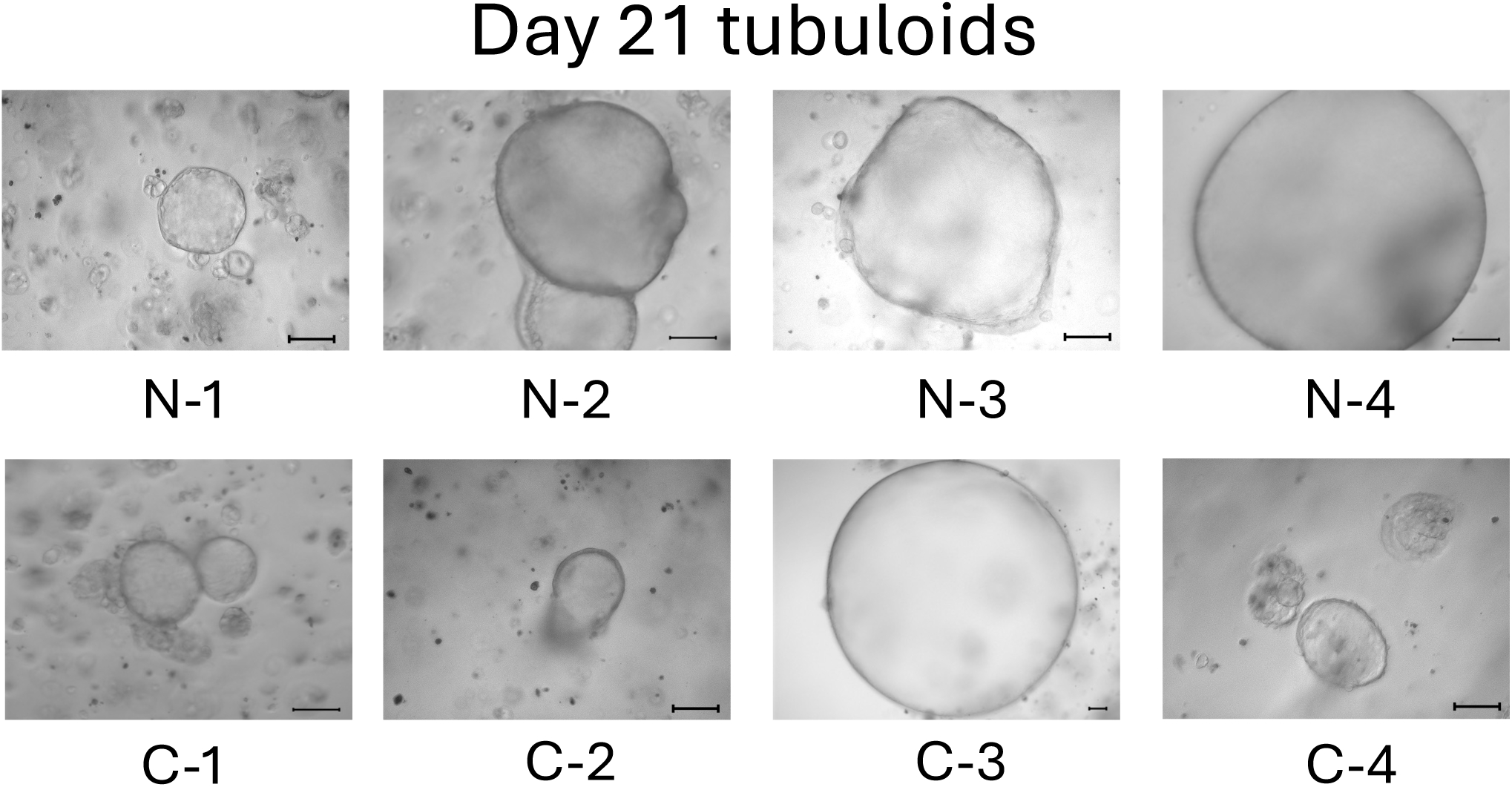
On day 21, all the tubuloids formed complete 3D structure. The top row shows 3D-cultured cells from individuals with kidney function (N-1 to N4), and the bottom row shows those from mild CKD (C-1 to C-4). Scale bars : 200µm

Our observations for 21 days after Matrigel administration revealed that tubuloids derived from normal cells grew faster overall, but with time, even mild CKD cell-derived tubuloids could produce beautifully rounded tubuloids that were comparable to those derived from normal cells.

## Discussion

Chronic kidney disease (CKD) is a disease composed of a wide variety of underlying causes, and its complexity makes it difficult to study. The additional difficulty is the fact that CKD is an aging and senescence-based diseases, and the most commonly used functional models of CKD are the mouse ones, whose average lifespan is approximately two years, making it difficult to accurately represent the gradual progression of CKD over the decades of human life. Another newly emerging technology, kidney iPSC organoids, are also difficult to represent human aging and senescence because they reflect fetal kidneys, although its sophistication is impressive.

The patient-derived organoids that we have successfully generated are likely to be ideal models for organ research, not only representing the current pathology of the individual, but also providing a true representation of aging and senescence, which is not possible with mice or iPSC organoids. As a case study that has already been reported, it has been found that renal tubular cysts shrink when tolvaptan is used in an adult organoid model of autosomal dominant polysistic kidney disease (ADPKD), but that tolvaptan does not work in an iPSC organoid-derived model of ADPKD [13].

Additionally, tubuloids can be formed by using a completely same protocol. In the case of kidney organoid, a completely same protocol cannot be used for inducing differentiation of iPSCs into nephron progenitor cells [13]. Usually, to compare the different genetic variants by using iPS kidney organoids, we need to use CRISPR/Cas9 system to produce the point mutation on the same iPSCs or embryonic stem cells (ESCs) [14, 15]. By this way, we can induce differentiation of iPSCs into nephron progenitor cells and kidney organoid by using a completely same protocol. Complete comparison between different individuals in a same condition is currently impossible by using iPS kindey organoids. Our tubuloids overcomes this matter. We can produce tubuloids from multiple individuals by using a completely same protocol and compare their response, although their developing phenotypes differ to some extent.

We have also recently succeeded in creating another CKD model by applying cisplatin to patient-derived tubuloids by accelerating cellular senescence [12]. Thus, patient-derived tubuloids as a research model for aging and senescence-based kidneys allow observations that are not possible in disease models such as rodents or iPSC organoids. Kidney tubuloids, which can be easily and rapidly produced in parallel in a same culture condition, are expected to play an active role in further elucidating pathological conditions and identifying new drugs in the future.

## Data Availability

All data produced in the present study are available upon reasonable request to the authors.

## Acknowledgement

We would like to thank the study participants who kindly allowed to give us the pieces of their resected kidneys.

## Data availability statements

Further information and requests for resources and reagents should be directed to and will be fulfilled by the Lead Contact, Yutaro Mori.

## Author contributions

M.M., Y.M. and Y.N. performed the experiments, collected and analyzed data, and wrote the manuscript. Y.M. and M.M. established the hRPTECs and Y.N. and S.M. helped the procedure. T.F., H.K., F.A., K.S., T.M., E.S., and S.U. supported the data analysis. Y.W., S.Y. and Y.F. resected the patients’ kidneys as standard treatment for malignant diseases. Y.M. developed experimental strategy, supervised the project, and edited the manuscript. All authors discussed the results and implications and commented on the manuscript.

